# Vertebrobasilar Dolichoectasia Future Aspects: A Meta-analysis of Clinical Features and Treatment Strategies

**DOI:** 10.1101/2024.12.06.24318631

**Authors:** Nour Shaheen, Santiago Ortega-Gutierrez, Edgar A Samaniego, Panagiotis Mastorakos, Michael Reid Gooch, Pascal Jabbour, Oliver Flouty, Kathleen Dlouhy, Mario Zanaty

**Affiliations:** Department of Neurosurgery, University of Iowa, Iowa City, IA, USA; Department of Neurology, Neurosurgery and Radiology, University of Iowa, Iowa City, IA, USA; Department of Neurosurgery, Thomas Jefferson University Hospital, Philadelphia, PA, USA; Department of Neurosurgery and Brain Repair, University of South Florida, Tampa, FL, USA

**Keywords:** vertebrobasilar dolichoectasia, dolichoectatic vertebrobasilar fusiform aneurysms, DVBFAs, endovascular, EVT, open surgery, conservative management, bypass, flow diversion

## Abstract

**Background:** Dolichoectatic Vertebrobasilar fusiform aneurysm (DVBFAs) presents a clinical challenge due to its complex anatomical features and associated neurological complications. This meta-analysis evaluates the clinical outcomes of endovascular treatment (EVT), open surgery, and conservative management for VBDA.

**Methods:** A systematic review of the PubMed, Scopus, and Web of Science databases was conducted to identify studies reporting on radiologically confirmed DVBFAs. Clinical outcomes were assessed using the modified Rankin Scale (mRS) and mortality rates. Meta-regression was performed to identify potential predictors of treatment outcomes.

**Results:** Ten studies with 219 patients were analyzed. Of the cohort, 58.4% underwent EVT, 24.6% received open surgery, and 16.9% were managed conservatively. The overall proportion of patients achieving a good clinical outcome (mRS <3) was 46.8%, with EVT showing the highest proportion at 59.4%, compared to 32.3% for open surgery and 24.7% for conservative management (p = 0.0145). The overall mortality rate was 25.98%, with EVT having the lowest mortality rate at 10.06%, followed by open surgery at 44.44% and conservative management at 63.30% (p = 0.0004). Subgroup analyses revealed statistically significant differences between treatment approaches in clinical outcomes and mortality.

**Conclusion:** EVT appears to provide better clinical outcomes for DVBFAs, though mortality rates remain high across all treatment modalities. The absence of significant differences in subgroup analysis suggests the need for further randomized controlled trials (RCTs) of EVT vs. conservative management to establish definitive treatment guidelines.

## Background

Dolichoectatic Vertebrobasilar) fusiform aneurysms (DVBFAs) are rare but serious conditions characterized by the elongation, dilation, and tortuosity of the vertebrobasilar arteries with aneurysmal formation ^1–6^. They are frequently associated with a range of neurological complications, including thromboembolism, brainstem compression, obstructive hydrocephalus, and, subarachnoid hemorrhage ^5,7–11^. Despite advancements in diagnostic imaging and treatment techniques, the optimal management of DVBFAs remains a significant clinical challenge ^12–15^. Treatment options generally include conservative management, endovascular therapy (EVT), and open cerebrovascular surgery, each of which carries its own risks and benefits ^16–22^.

The current body of evidence lacks a comprehensive review that evaluates the clinical outcomes of different treatment approaches for DVBFAs. Furthermore, there remains significant uncertainty regarding the association of various clinical risk factors—such as hypertension, diabetes, and atherosclerosis—with treatment outcomes, making it difficult to predict which patients are more likely to benefit from specific interventions. To address these gaps, we conducted a systematic meta-analysis of available studies on DVBFAs, with a particular focus on the effectiveness of EVT, open surgery, and conservative management. In addition, we aimed to explore potential predictors of treatment success and assess the sample size requirements for future randomized controlled trials (RCTs).

## Methods

### Protocol and registration

We carried out this systematic review in compliance with the Preferred Reporting Items for Systematic Reviews and Meta-Analyses (PRISMA) guidelines ^23^. The detailed PRISMA 2020 checklist can be found in the supplementary material. Additionally, we registered the review protocol in advance with PROSPERO (registration ID: CRD42024589232).

### Literature Search

We searched the PubMed, Scopus and Web of Science databases from their inception until July 18, 2024. Using Boolean operators “OR” and “AND,” we employed medical subject headings (MeSH) terms and keywords: “dolichoectasia” OR “dolichoectasis” OR “dolichoectatic” AND “vertebral artery” OR “basilar artery” OR “vertebrobasilar” OR “internal cerebral artery” AND “fusiform aneurysms’NOT “saccular aneurysm” OR “blister aneurysm” (Supplementary Table 1)). All articles were uploaded to EndNote, where duplicates were subsequently removed.

### Study Selection

Included studies met the following criteria: 1) involved patients with radiologically confirmed DVBFAs, including the vertebral artery, basilar artery, or vertebrobasilar artery; 2) provided data on clinical features, treatment protocols, and outcomes; and 3) were written in English. Excluded studies were those that 1) were literature reviews, case reports, editorials, book chapters, technical notes, abstracts, or autopsy reports; 2) did not clearly differentiate data of patients with DVBFAs from those with other types of aneurysms; 3) did not specify the involved artery; 4) focused on saccular aneurysms, blister aneurysms, fusiform ICA aneurysms, fusiform vertebral artery aneurysms, fusiform basilar artery aneurysms, or vertebrobasilar fusiform aneurysms; or 5) had significant data insufficiencies, such as missing clinical characteristics or treatment outcomes.

Two authors (Nour Shaheen and Mario Zanaty) independently reviewed the titles and abstracts of all extracted citations and then evaluated the full texts of articles that met the inclusion criteria. Another author resolved any disagreements. Eligible studies were included, and references were screened to identify additional relevant studies.

### Data Extraction

We developed a Python script to automate the extraction of key data on DVBFAs from selected articles. The script begins by setting up the environment and installing essential libraries for handling PDFs, interacting with Google’s Gemini AI, and managing the user interface. We utilized a function called ‘extract_variable()’, which uses tailored prompts and Gemini AI to extract specific information such as study ID, design, age, sex, country, total patient number, risk factors, and clinical data from each paper. Missing data were input as NA. To ensure reliable variable extraction, the articles were systematically reviewed, followed by automated extraction, which was then carefully verified through manual review. The extracted data were then compiled into a table, saved as a CSV file, and prepared for meta-analysis.

### Data Synthesis and Quality Assessment

We used the modified Rankin Scale (mRS) to evaluate the clinical outcomes of different treatment groups (EVT, surgery and conservative) after a follow-up period of at least three months. The mRS scores post-intervention were compared with pre-intervention scores. A score of ≤ 2 on the mRS was deemed a favorable\good outcome, whereas a score of ≥ 3 was considered unfavorable\poor. Mortality rates were also considered as secondary outcomes. We tracked changes in mRS from admission to the most recent follow-up.

The methodological quality of the included studies was assessed using the Oxford Centre for Evidence-Based Medicine (OCEBM) Levels of Evidence (Levels I-V) ^24^. No studies were excluded based on quality, but study quality was taken into consideration when interpreting the results.

### Statistical Analysis

All statistical analyses were conducted using R (version R-4.4.1, The R Foundation for Statistical Computing) and RStudio (Version 2024.04.2, RStudio, Inc.). The “meta” package was used for meta-analyses, and ggplot2 was used for data visualization.

Continuous variables were described using odds ratio (OR) and 95% confidence intervals. Categorical variables were presented as frequencies and percentages or as weighted proportions (proportion effect sizes) with 95% confidence intervals. The chi-square test was employed to assess the significance between pre-and post-mRS scores.

Funnel plots and Egger’s test (where a p-value < 0.05 suggests bias) were employed to detect publication bias. We verified the visual symmetry of all studies, and endpoints with a sufficient number of articles (n ≥ 9) were assessed using Egger’s test ^25^, No publication bias was detected in any of the endpoints tested.

We performed subgroup analyses to evaluate statistical variations across different groups. Statistical significance was determined by two-tailed p-values less than 0.05. To examine heterogeneity, we applied the chi-square test along with Higgins’ I² test ^26^. We used **random-**effects models for all meta-analyses due to anticipated heterogeneity across studies. Heterogeneity was assessed using Cochran’s Q test and Higgins’ I² test, with I² values above 50% indicating substantial heterogeneity. We accounted for heterogeneity by exploring potential sources through subgroup and sensitivity analyses.

For sensitivity analysis, we conducted a “leave-one-out” analysis to evaluate the robustness of the pooled results. This approach involved systematically excluding one study at a time to ensure that no single study disproportionately influenced the overall effect size.

#### Meta-Regression Analysis

We performed a meta-regression analysis to investigate the relationship between treatment outcomes and potential predictors, such as age, follow-up duration, symptomatic status, hypertension, diabetes, and atherosclerosis. Predictors were selected based on clinical relevance and availability of data in the included studies. To control for multiple comparisons, we applied Bonferroni correction when examining multiple variables in the meta-regression analysis.

#### Sample Size Calculation

We calculated the required sample size for comparing two treatment groups (e.g., EVT vs. Conservative) based on ORs. We first conducted a random-effects meta-analysis to combine effect sizes effect size (h) across studies. To estimate the effect size, we calculate Cohen’s *h*. Next, We computed the log-transformed odds ratios and their variances. Finally, we perform power analysis using the equation: N= [((Zα/2 +Zβ) ² × 2 × B²) / d²] to determine the required sample size, based on the effect size, a significance level of (α = 0.05), and a Power (1 − β) of 0.95.

## Results

### Study Selection and Quality Assessment Results

An initial search of the PubMed, Scopus, and Web of Science databases identified 1,161 articles (Fig. 1). After removing duplicates, 798 unique articles remained. Screening titles and abstracts led to the exclusion of 708 studies. We retrieved and assessed 90 papers for potential inclusion. Of these, 80 articles did not meet our inclusion criteria and were excluded. 10 articles met the inclusion criteria and were incorporated into the final analysis. All included studies were classified as Level III according to the OCEBM Levels of Evidence ^16–22,27–29^. (Figure 1)

**Figure 1.**
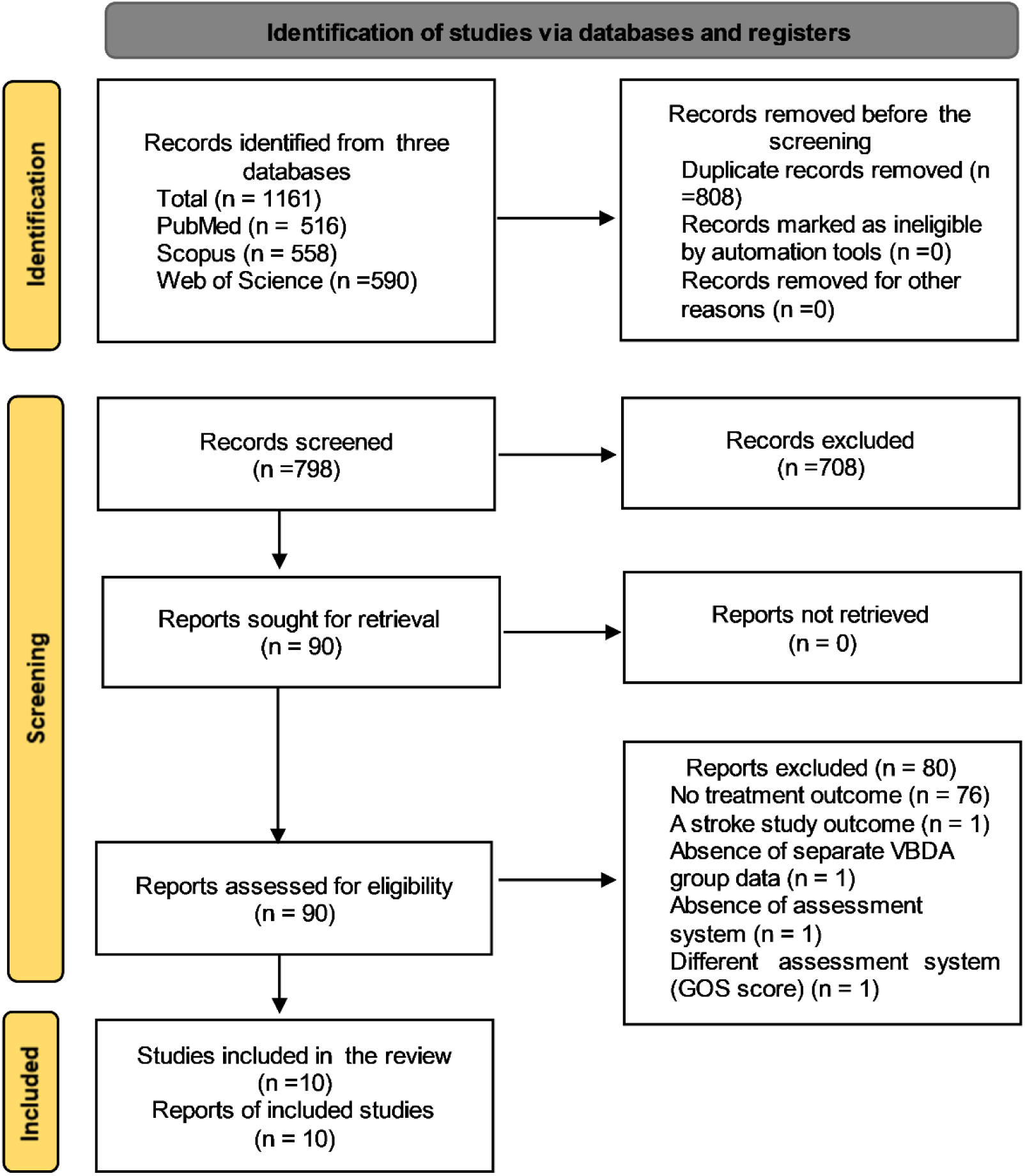
PRISMA flow diagram of studies’ screening and selection.

### Baseline study characteristics

The pooled cohort included 219 patients with a mean age of **58.91 years** (SD = 4.01), and a male predominance (77.6%). Of these, 128(**58.4%)** received EVT, 54(**24.6%)** underwent open surgery, and 37(**16.9%)** were managed conservatively. The mean follow-up duration was **31.17 months** (SD = 19.24). Symptomatic presentation was noted in **45.6%** of patients, and common clinical features included brainstem compression in **28.3%**, ischemic symptoms in **9.6%**, and hypertension in **28.3%** of the cohort.

Arterial aneurysms were located in the basilar and vertebral arteries in 111 patients, in the basilar artery alone in 102 patients, and in the vertebral arteries alone in the remaining 2 cases. The mean length of DVBFAs was 28.05 mm (SD = 6.14 mm), and the mean diameter was 14.72 mm (SD = 1.38 mm). Regarding risk factors are presented in Table 1.

**Table 1.** Summary of studies evaluating treatment approaches for DVBFAs across various locations including study design, patient demographics, treatment types, aneurysm locations, and mortality rates.

| Study ID | location | Study design | Total Study Size | Treatment | Group No | Mean age (SD) | Male, N (%) | Follow up(m) | Aneurysm location |  |  | Mortality rate N (%) |
| --- | --- | --- | --- | --- | --- | --- | --- | --- | --- | --- | --- | --- |
|  |  |  |  |  |  |  |  |  | BA | VA | BA+VA |  |
| Ji Z 2024<sup>20</sup> | United States | single-centre retrospective study | 47 | EVT (flow diversion (FD)) | 22 | 59.4 (8.7) | 17 (77.3%) | 23 |  |  | 22 | 2 (9.1%) |
|  |  |  |  | EVT (stenting with low-profile braided stents (LPBS)) | 25 | 58.6 (10.2) | 21 (84%) | 28 |  |  | 25 | 3 (12%) |
| Siddiqui AH 2023<sup>21</sup> | United States | Retrospective study | 27 | EVT + Triple therapy | 14 | 60.8(2.4) | 12 (85.7%) | 38.8(40.3) | 12 |  | 2 | 0 |
|  |  |  |  | EVT + DAPT | 13 | 60.6(15.7) | 9 (69.2%) | 38.8(70.04) | 9 |  | 4 | 1(7%) |
| Rezai Jahromi B 2022<sup>22</sup> | Japan | Retrospective Case Series | 6 | Open (Slow-Closing Clip) | 6 | 60(8.6) | 5 | 57.6(55.9) | 6 |  |  | 3(50%) |
| Nakatomi H 2020<sup>16</sup> | United States | Retrospective multicenter cohort study | 32 | Open | 21 | 56.8 | 12 | 56.1 (60.8) | 11 |  | 10 | 12(57%) |
|  |  |  |  | Conservative | 11 |  | 8 | 19.4 (26.7) | 7 |  | 4 | 10(90%) |
| He X 2019<sup>19</sup> | England | Retrospective study | 19 | EVT (stenting with and without coils) | 19 | 52(6.25) | 17 (89.5%) | 5.6(4.03) |  |  | 19 | 1(5%) |
| Wang J 2019<sup>27</sup> | Switzerland | single-centre retrospective study | 22 | EVT (stenting with and without coils) | 22 | 52.6 (12.6) | 22 (100%) | 49.6 (27.7) | 4 |  | 18 | 7(31%) |
| <sup>28</sup>N 2018 | Japan | Retrospective Study | 4 | Open | 4 | 67.5 (4.5) | 3 (75.0%) | 6 | 4 |  |  | 2 (50.0%) |
| Zhang H 2018<sup>29</sup> | United States | single-centre retrospective study | 12 | EVT (stenting with coils) | 4 | 63.2(5.8) | 3(75%) | 19.7(4.03) | 3 |  | 1 | 0 |
|  |  |  |  | Open | 7 | 53.7(4.8) | 6(85%) | 19.2(21.5) | 4 | 2 | 1 | 228%) |
|  |  |  |  | Conservative | 1 | 59 | 0 | 7 | 1 |  |  | 0 |
| Lawton MT 2016<sup>17</sup> | United States | Retrospective study | 41 | Open | 16 | 59 | 12(75.0%) | 64.8 | 16 |  |  | 5 (20%) |
|  |  |  |  | Conservative | 25 | 63 | 16 | 44.4 (3-140) | 25 |  |  | 12(48%) |
| Wu X 2013<sup>18</sup> | United States | Retrospective study | 9 | EVT (stenting with coils) | 9 | 59.56(11.5) | 7(77.8%) | 20.7 (6.9) |  |  | 9 | 1(11%) |

### Clinical Characteristics by Treatment Group

The EVT group included 128 patients with a mean age of 58.1 years (SD = 3.9), while the open group had 54 patients with a mean age of 60.8 years (SD = 5.1), and the conservative group had 37 patients with a mean age of 59.6 years (SD = 3.1). The follow-up duration was 10.7 months (SD = 13.8) for the EVT group, 22.5 months (SD = 26.3) for the open group, and 23.6 months (SD = 19.0) for the conservative group.

Regarding presenting clinical features, brainstem compression was present in 62 patients (46%) in the EVT group, while none in the open or conservative groups had this condition. Cerebral infarctions around the DVBFAs territory were noted in 19 patients (14.8%) in the EVT group, 4 patients (7.4%) in the open group, and none in the conservative group. The prevalence of symptomatic patients was 54 (42.2%)in the EVT group, 34 (63%)in the open group, and 12 (32.4%)in the conservative group. Hypertension was significantly more common in the EVT group 52 (40.6%)compared to the open 9 (16.7%)and conservative groups 1 (2.7%). (Supplementary Table 2S)

### Results of Meta-Analysis

#### I. Proportion analysis of mRS scores after the treatment

##### a) Good clinical outcome: mRS <3

Across all treatments, the overall proportion of patients achieving a good clinical outcome (mRS <3) was 46.8% (95% CI: 34.2% to 59.8%), EVT demonstrated the highest proportion of favorable outcomes, with 59.4% (95% CI: 50.7% to 67.5%) of patients achieving mRS <3, compared to 32.3% (95% CI: 17.0% to 52.6%) for open surgery and 24.7% (95% CI: 6.6% to 60.5%) for conservative management (p = 0.0145 for subgroup differences). Moderate heterogeneity was observed across studies (τ² = 0.4909, I² = 28.8%) (Figure 2a), suggesting some variability in treatment effects on clinical outcomes (Figure 2a).

**Figure 2.**
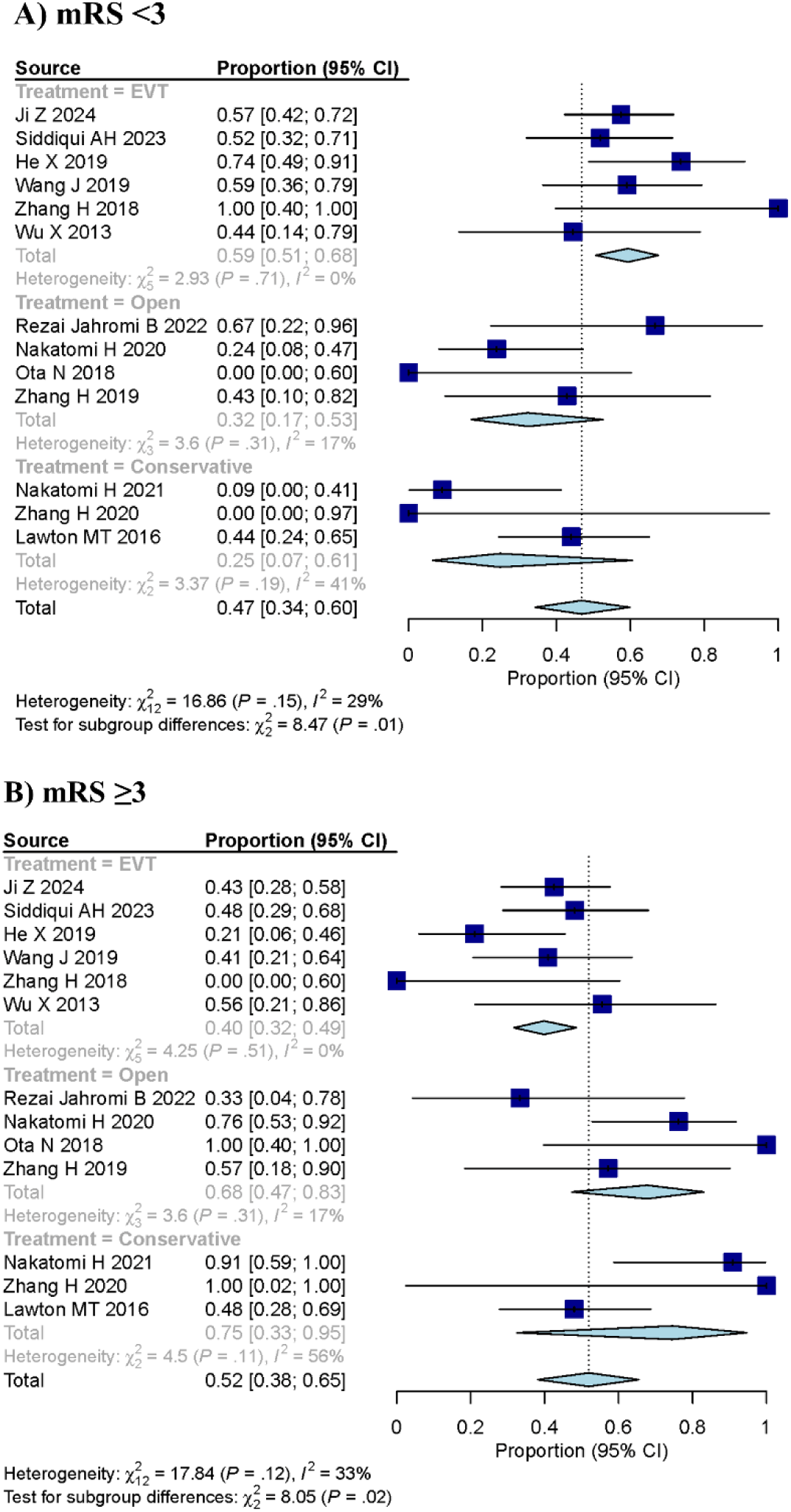
Forest plot showing proportion analysis of a) mRS<3 and b) ≥3 scores after the treatment

##### b) Poor clinical outcome: mRS ≥3

The overall proportion of patients with poor clinical outcomes (mRS ≥3) was 51.95% (95% CI: 38.13% to 65.47%). EVT was associated with a significantly lower proportion of poor clinical outcomes (39.84%, 95% CI: 31.74% to 48.55%) compared to open surgery (67.71%, 95% CI: 47.38% to 83.00%) and conservative management (74.54%, 95% CI: 32.51% to 94.68%) (p = 0.0179). Moderate heterogeneity was noted (τ² = 0.5983, I² = 32.8%, H = 1.22), suggesting some variability across studies. The test for subgroup differences was statistically significant (p = 0.0179), emphasizing the impact of treatment modality on clinical outcomes (Figure 2b).

#### II. Treatment Mortality

The overall mortality rate across all treatments was 25.98% (95% CI: 13.36% to 44.41%). EVT demonstrated the lowest mortality rate at 10.06% (95% CI: 4.34% to 21.63%), followed by open surgery at 44.44% (95% CI: 31.87% to 57.78%) and conservative management at 63.30% (95% CI: 26.87% to 89.01%) (p = 0.0004). There was high heterogeneity among the studies for mortality (τ² = 1.6892, I² = 66.3%), suggesting that treatment effects vary significantly across studies (Supplementary Figure 1S).

#### III. Comparison of mRS scores pre- vs post-treatment

##### a) Good clinical outcome at presentation: mRS <3

In the comparison of mRS scores pre- and post-intervention, EVT was associated with a greater likelihood of achieving good outcomes post-treatment, with an OR of 2.22 (95% CI: 0.71–6.97). Open surgery had an OR of 1.46 (95% CI: 0.48–4.41), and conservative treatment showed the largest increase in odds for good outcomes (OR 2.77, 95% CI: 0.93–8.25). However, the test for subgroup differences was not statistically significant (p = 0.7149), indicating no evidence of treatment superiority across the three approaches. The overall random-effects model showed a pooled OR of 2.13 (95% CI: 1.06–4.28; p = 0.0347), suggesting a statistically significant association between interventions and good outcomes. Heterogeneity across studies was moderate (τ² = 0.5753; I² = 46.6%, p = 0.0511). (Figure 3a)

**Figure 3.**
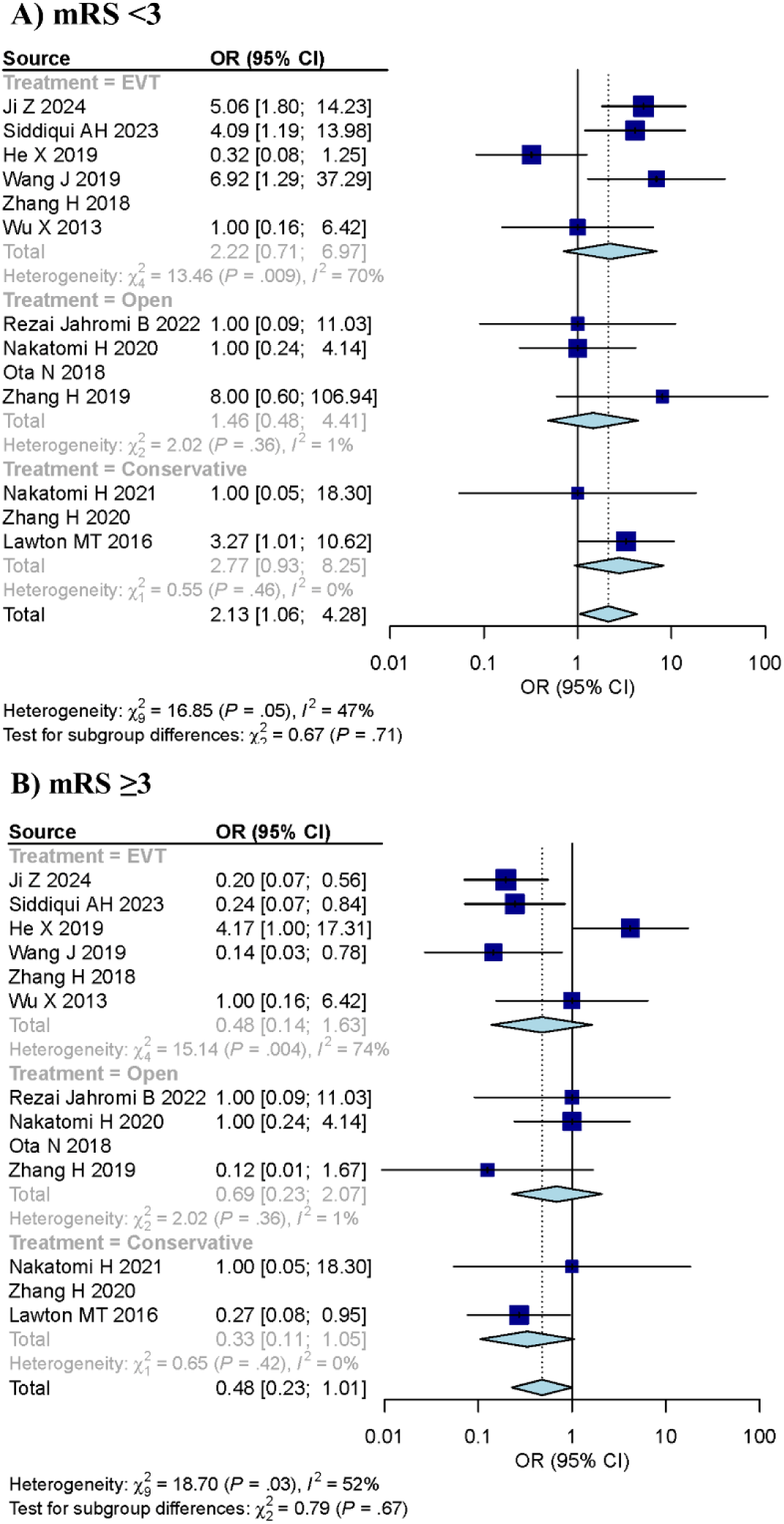
Forest plot showing a) mRS<3 and b) ≥3 scores pre- versus post-treatment

##### b) Poor clinical outcome at presentation: mRS ≥3

For patients with an mRS ≥3 at presentation, EVT demonstrated a trend toward reducing the likelihood of poor outcomes, with an OR of 0.48 (95% CI: 0.14–1.63). Open surgery showed a similar trend (OR 0.69, 95% CI: 0.23–2.07), while conservative treatment had the largest reduction in poor outcomes (OR 0.33, 95% CI: 0.11–1.05). The test for subgroup differences was not statistically significant (p = 0.6745), providing no evidence that any one treatment approach is superior to the others in reducing poor clinical outcomes. The overall random-effects model showed an OR of 0.48 (95% CI: 0.23–1.01; p = 0.0530), and heterogeneity across studies was moderate (τ² = 0.7159; I² = 51.9%, p = 0.0278) (Figure 3b).

### Risk of bias assessment

The funnel plot assessment for mRS <3, using the linear regression test (t = −1.14, df = 7, p = 0.2909), did not reveal significant evidence of publication bias, with a bias estimate of −2.11 (SE = 1.84). Similarly, the funnel plot assessment for mRS **≥3**, based on the linear regression test (t = 1.10, df = 7, p = 0.3061), also did not indicate significant evidence of publication bias, with a bias estimate of 2.20 (SE = 1.99) (supplementary Figures 2S & 3S).

### Sensitivity Analysis

The **sensitivity analysis**, conducted by omitting one study at a time, showed that the pooled OR remained consistent across all studies, further supporting the robustness of the findings (supplementary Figures 4S & 5S).

### Meta-regression analysis

In the univariate meta-regression analysis, none of the predictors (age, follow-up duration, symptomatic status, and clinical factors such as hypertension, diabetes, and atherosclerosis) showed a statistically significant association with outcomes. Additional predictors, including headache pres ence, infarction, vascular compression, hemorrhagic history, and ischemic status, also did not sho w a statistically significant association with treatment outcomes. This suggests that these factors did not strongly influence the effectiveness of the treatments analyzed (Supplementary Figure 5 & 6S and Table 3 & 4S).

### Sample Size and Effect Size Calculation

Based on the sample size calculation using arcsine transformation, approximately **18** participants per group would be needed to detect an effect size (h) of 0.84, with a significance level of 0.05 and a power of 0.95. This calculation indicates that future trials comparing EVT and conservative management should have a minimum of 9 patients in each group to achieve statistical power.

## Discussion

The meta-regression analysis reveals that the type of presentation (hemorrhagic vs. ischemic) and symptom status (symptomatic vs. asymptomatic) were not significantly associated with the clinical outcome. Only the type of treatment predicted the clinical outcome We have found that EVT is associated with significantly better clinical outcomes (mRS <3) and lower mortality compared to open surgery and conservative treatment in patients with DVBFAs regardless of their mRS at presentation. While EVT showed a clear advantage in the overall proportion analysis, the subgroup analysis did not reveal statistically significant differences between treatment modalities. This highlights the complex nature of managing DVBFAs. The discrepancies between the proportion and subgroup analyses could be attributed to small sample sizes, wide confidence intervals, and variability in patient populations, leading to potential underpowering in the subgroup analysis. However, the consistency in effect sizes, indicated by low heterogeneity in the proportion analysis, supports the reliability of these findings. Thus, we believe the proportion analysis provides a more robust and reliable estimate of EVT’s relative effectiveness in this context. Despite this, conservative management remains a valid approach until large-scale RCTs can directly compare EVT and conservative treatment to definitively establish the optimal management strategy for DVBFAs.

^7,16^. A recent meta-analysis examining the natural history of VBDE, involving 827 patients over 5,093 patient years, found an annual mortality rate of 13% ^30^. Despite advancements in endovascular techniques, such as the use of flow diverters, and improved surgical revascularization skills, patient outcomes remain suboptimal, with both EVT and surgery reporting high mortality rates (54%–100%) and significant morbidity rates (15%–26%) ^16,17,27,31^.

Our findings align with previous reports that highlighted the high mortality rates among patients treated conservatively ^8,32–35^. However, our results suggest that EVT offers more favorable outcomes, contrasting with earlier studies that reported poor results across all treatment approaches ^12–14^. The improved outcomes observed with EVT may be due to advancements in endovascular techniques, better antiplatelet management (possible triple therapy), and more refined careful planning, which were not available in previous research. There was treatment variability in each arm. The endovascular group used closed-cell stents or flow diversion while some added oral anticoagulation on top of dual antiplatelet therapy. There is also a difference in the number of stents deemed necessary to provide adequate flow diversion and whether the non-implanted vertebral artery was sacrificed. Conservative management included triple therapy, dual antiplatelet therapy and in some instances other drugs such as tetracycline and angiotensin receptor blockers. Similarly, open surgery treatment varied between clipping, parent and parent artery occlusion, with or without bypass. This complex planning and heterogeneity in treatment makes any meta-analysis difficult to perform. Unfortunately, further subdivision of each treatment arm will render the number of patients too small for meaningful statistical inference.

Although there were some differences between groups, such as a higher incidence of brainstem compression in the EVT group. We do not have an explanation on why brainstem compression is not reported in surgical case series. It could be most likely that it was present but not noted in the published manuscripts. Another explanation would be that advanced cases are referred for endovascular if surgery is deemed futile or extremely risky. This infers a selection bias. Thus, we chose to assess patients using mRS scores, which could reflect underlying compression. The mRS is a well-established numerical measure of disability and functional outcomes, making it a reliable criterion for comparing treatment efficacy, particularly in RCTs. By focusing on mRS, we aimed to standardize outcome assessments, reducing potential biases arising from variability in patient presentation and baseline characteristics.

Despite the heterogeneity observed in the literature, our sensitivity analysis demonstrated that the results were robust to the omission of individual studies. This robustness strengthens the reliability of our findings, indicating that the overall conclusions remain consistent despite variations in study design, patient populations, and treatment protocols. The consistent results across different analytical scenarios enhance the credibility of our conclusions.

The meta-regression analysis revealed that none of the predictors (ageBA involvement, VBA involvement) or clinical factors (hypertension, diabetes, and atherosclerosis) were significantly associated with the outcomes. While this may be due to the small sample size, it underscores the need to identify biomarkers that can better distinguish between patients who are likely to experience good versus poor outcomes. Notably, some predictors, such as hemorrhagic history and infarction, displayed higher coefficient values, hinting at possible, albeit statistically non-significant, associations. However, these findings suggest that these factors alone are unlikely to drive substantial differences in treatment efficacy.

Future research should focus on conducting RCT comparing EVT and conservative treatment to establish definitive treatment guidelines for VBDA. Additionally, identifying biomarkers that predict treatment success could improve patient selection and outcome prediction. Standardizing EVT protocols would also help reduce variability in clinical outcomes.

In clinical practice, EVT may offer a promising treatment option for patients with VBDA, especially those at high risk, given its association with improved outcomes and reduced mortality. However, further research is required to optimize treatment strategies and ensure better outcomes for all patients.

### Limitations

Despite these promising results, several limitations should be acknowledged. First, the majority of the included studies were retrospective, increasing the risk of selection bias and limiting control over confounding factors. The relatively small sample sizes, particularly in the subgroup analyses, further reduced the statistical power, potentially leading to underpowered conclusions. Additionally, the high heterogeneity in mortality outcomes (I² = 66.3%) suggests significant variability in treatment protocols and patient populations, which complicates direct comparisons. While random-effect models were employed to account for this variability, the findings should be interpreted with caution.

The absence of predefined subgroups in the included studies may also have contributed to the lack of significant findings in subgroup analyses. Furthermore, variability in follow-up durations could have influenced assessments of long-term outcomes, such as mortality and functional recovery. The lack of standardized treatment protocols and consistent definitions of VBDA across studies adds to this variability, limiting the generalizability of the findings.

Another limitation is the absence of RCT, which weakens the strength of the evidence. Future RCTs are needed to confirm these findings and establish clear treatment guidelines. Although no strong evidence of publication bias was detected, it is possible that negative or inconclusive studies were underreported, potentially skewing the results. Lastly, the predominance of studies conducted in regions like the United States and Japan may limit the applicability of the findings to other populations, further highlighting the need for well-designed, prospective trials.

## Conclusion

Our meta-analysis suggests that EVT may be associated with more favorable clinical outcomes and reduced mortality, showing promise as an effective treatment for improving outcomes in patients with DVBFAs. However, the mixed results—particularly the lack of statistical significance in subgroup analyses—suggest that conservative management remains a valid approach until large-scale RCTs directly compare EVT with conservative treatment. Future research should prioritize identifying predictors of treatment success and refining optimal management strategies for DVBFAs.

## Data Availability

All data, analytic code, and materials used in this review are available upon reasonable request ?from the corresponding author.?

## Availability of Data, Code, and Other Materials

All data, analytic code, and materials used in this review are available upon reasonable request from the corresponding author.

